# Safety and efficacy of antiviral therapy alone or in combination in COVID-19 - a randomized controlled trial (SEV COVID Trial)

**DOI:** 10.1101/2021.06.06.21258091

**Authors:** SEV COVID trial group, Budha O Singh, Bikram Moirangthem, Prasan Kumar Panda, Yogesh Arvind Bahurupi, Sarama Saha, Girraj Saini, Minakshi Dhar, Mukesh Bairwa, Venkatesh Srinivasa Pai, Ankit Agarwal, Girish Sindhwani, Shailendra Handu, Ravikant

## Abstract

**Background:** Definitive antiviral treatment is not available for COVID-19 infection except remdesivir that even with many doubts. Various combination antivirals have been tried.

**Methods:** A single-center, open-label, parallel-arm, stratified randomized controlled trial evaluated the therapeutic potential of hydroxychloroquine and lopinavir-ritonavir in combination with ribavirin in COVID-19. Enrolled patients in severe category were randomized into three groups: A: standard treatment, B: hydroxychloroquine+ribavirin+standard treatment, or C: lopinavir+ritonavir+ribavirin+standard treatment; while non-severe category into two groups: A: standard treatment or B: hydroxychloroquine+ribavirin. Combination antivirals was given for 10 days and followed for 28 days. The primary endpoints were safety, symptomatic and laboratory recovery of organ dysfunctions, and time to SARS-CoV-2 RT-PCR negative report.

**Results:** Total 111 patients randomized: 24, 23, and 24 in severe category A, B, and C respectively, and 20 in each non-severe group. Two patients receiving ribavirin experienced drug induced liver injury and another developed QT prolongation after hydroxychloroquine. In the severe category, 47.6%, 55%, and 30.09% in A, B, and C groups respectively showed symptomatic recovery compared to 93.3% and 86.7% in A and B groups respectively in the non-severe category at 72hrs (P>0.05).

**Conclusions:** The results failed to show statistical superiority of the antiviral combination therapies to that of the standard therapy in both the severe and non-severe categories in symptomatic adult patients of COVID-19. However, results do indicate the benefit of non-standard interventional combination therapy in severe disease. Furthermore, the dose of ribavirin needs to be reconsidered in the Indian population.

## BACKGROUND

COVID-19 is an infectious disease caused by the novel coronavirus SARS-CoV-2. Individuals of all age groups and both sexes are at risk of infection with a higher probability of severe disease in the elderly, pregnant, transplant recipients, and people with chronic medical conditions(1)(2)(3). The disease can progress from asymptomatic to mild cough, fever, and sore throat to acute respiratory distress syndrome, various other end-organ involvements like cardiac, dermatologic, hematological, hepatic, neurological, and renal, thromboembolic events, septic shock, or death, or even long COVID(4)(5)(6)(7)(8)(9). These presentations can be categorized as non-severe and severe COVID-19 during disease progression from the viremia phase to the immunological phase.

Antiviral therapies are expected to have a higher benefit if administered early in the course with immunosuppressive or anti-inflammatory therapies which are likely to be beneficial in the later stages of COVID-19 infection. Supportive treatment forms the mainstay of non-severe cases of COVID-19 infections. Hydroxychloroquine (HCQ) is thought to be effective in treatment and prophylaxis(10)(11). It has got immunomodulatory effects and it may also interfere with the binding of severe acute respiratory syndrome-associated coronavirus (SARS-CoV) to the cell receptor thus inhibiting viral entry(12).

The replication of severe acute respiratory syndrome coronavirus 2 (SARS-CoV-2) depends on two proteases viz. 3-chymotrypsin-like protease (3CLpro) and papain-like protease (PLpro) are responsible for cleaving polyproteins into an RNA-dependent RNA polymerase and a helicase. Lopinavir-ritonavir combination has also been tried in hospitalized patients due to its probable mechanism of inhibiting these proteases (13) (14). Ribavirin on the other hand has shown to have direct antiviral activity in vitro against the SARS virus and any strain variants that may emerge (15). The numerous studies on these individual antivirals and HCQ had failed to establish any significant benefits. However, combination therapy of these has not been attempted. Hence our study aims to define the outcome with these agents in combination in comparison to that of the standard supportive therapy.

## Methods

### STUDY SETTINGS, DESIGN, AND POPULATION

The study was a single-center, open-label, parallel-arm, stratified randomized controlled trial conducted at All India Institute of Medical Sciences (AIIMS), Rishikesh, Uttarakhand, India between March 2020 to October 2020 after obtaining approval of Institutional Ethics Committee. The trial was registered at the Clinical Trial Registry of India (CTRI/2020/06/025575).

Being an exploratory trial, in a relatively new infection, all consenting patients ≥18 years old, diagnosed with symptomatic COVID-19 disease with a positive reverse-transcriptase– polymerase chain reaction (RT-PCR) assay for SARS-CoV-2, were included in the trial. Exclusion criteria included: patients on medications which were contraindicated with lopinavir/ritonavir, hydroxychloroquine/chloroquine, or ribavirin; patients taking lopinavir-ritonavir based anti-retroviral therapy or on hydroxychloroquine/chloroquine or ribavirin; known allergic reactions to any of the drugs used in the treatment arms; inability to take oral medications (lopinavir-ritonavir, hydroxychloroquine/ chloroquine, ribavirin); pregnant or lactating females; patients who had received any of the experimental therapies for 2019-nCoV (off-label, compassionate use, or trial-related) within 30 days before participation in the present study.

## INTERVENTION

The enrolled patients were categorized as “severe disease” or “non-severe disease”, based on pre-defined criteria (WHO classification of COVID-19 severity). “Severe disease” was defined as confirmed pneumonia on chest imaging, SpO2 <93%, PaO2:FiO2 <300, respiratory failure (need for mechanical ventilation), septic shock, multiple organ dysfunction syndromes, liver disease (Child-Pugh score ≥ C, AST >5 times upper limit), renal impairment (estimated glomerular filtration rate ≤30 mL/min/1.73 m2) or receiving renal replacement therapy (continuous renal replacement therapy, hemodialysis, or peritoneal dialysis), and other single organ failures (e.g., heart, if specific definitions were met (NYHA classification for heart failure). The “non-severe disease” category included mild to moderate disease not fulfilling the criteria for severe disease.

Enrolled patients in the “severe disease” category were randomized dynamically into three arms: severe A, severe B, severe C. Severe A received standard treatment as per institute protocol. Severe group B received a combination of HCQ + ribavirin + standard treatment. Severe group C received lopinavir + ritonavir + ribavirin + standard treatment; while “non-severe disease” category patients were randomized into two arms: non-severe A and B. Non-severe A received the standard treatment and non-severe B received HCQ + ribavirin + standard treatment.

At the time of enrolment, demographic data were collected from the participants, including relevant data on their medical history, co-morbidities, as well as risk factors for severe COVID-19. Baseline investigations included a complete blood count, liver function tests, kidney function tests, glucose-6-phosphate dehydrogenase levels, and electrocardiography. Patients were followed up for 28 days from the day of enrolment.

### RANDOMISATION

The randomization was done with a computer-based software “randomize R package” of version 1.4.2. Randomization was done in blocks of four and patients were stratified into non-severe and severe as per defined criteria of severity.

### COMPARATOR

In the severe category three parallel arms of interventions were compared while two in the non-severe category.

## OUTCOMES/ENDPOINTS

The primary endpoints were safety, clinical recovery of symptoms and laboratory recovery of each organ involvement, and time to SARS-CoV-2 RT-PCR negative report of nasopharyngeal/throat swab specimen. Safety was determined by observing the frequency and severity of serious adverse events as per the division of acquired immune deficiency syndrome (DAIDS) table for grading the severity of adverse events. Clinical recovery was defined as normalization of fever, respiratory rate, oxygen saturation, and alleviation of cough at first follow-up at 72hrs of initiation of therapy, all sustained for at least 72 hours. Laboratory recovery was also applied at 72hrs intervals. Normalization and alleviation criteria were: fever – axillary temperature ≤36.9°C, oral temperature ≤37.2 °C, respiratory rate ≤24/minute on room air, SpO2 >94% on room air, mild or absent cough using a patient-reported ordinal scale (severe, moderate, mild, or absent).

The secondary endpoints included all-cause mortality, respiratory progression (defined as SpO2 ≤ 94% on room air or PaO2/FiO2 <300 mmHg and requirement for supplemental oxygen or more advanced ventilator support such as non-invasive/invasive ventilation), and hospital duration of stay.

Patients were assessed for clinical and laboratory improvements at 72 hours of initiation of treatment, same repeated once every 72 hours until the primary endpoint was met. Investigations included complete blood counts, liver function tests, renal function tests, HbA1c, blood glucose, PT/INR, serum electrolytes, arterial blood gases, chest X-ray, ECG, other organ markers as per involvement, and RT-PCR for SARS-COV-2 (until negative on two occasions at least 24 hours apart). Patients who failed to respond to the current treatment arm were shifted to treatment as per the institute protocol and as per the prevailing treatment guidelines. All adverse events were recorded at the first instance including dates as appropriate. Each adverse event was assessed for severity, causality, seriousness, and expectedness. If any serious adverse event was recorded, the participant was withdrawn from the trial, and management was done accordingly as per institute protocol.

## STATISTICAL ANALYSIS

Data were described as mean ± SD and proportions. Comparisons of the categorical variables of the arms were performed using chi-square or Fisher’s exact test. Participants enrolled in severe strata were analyzed using One-way ANOVA followed by a post hoc test. An independent t-test was applied for the comparison of means between two arms in non-severe strata. We followed per-protocol analysis for the entire study.

## Results

In the study, a total of 550 patients were screened and of which 111 patients were enrolled after randomization (Fig. 1). 71 participants were enrolled in the severe category and of which 24, 23 and 24 participants were randomized in groups A, B, and C respectively. 40 participants were categorized into the non-severe category, 20 participants in each group A and B. Two participants: one from the severe B and one from non-severe B showed an adverse drug reaction in the form of elevated liver enzymes (after receiving 2.4gm ribavirin per oral stat followed by 1.2gm twice daily) which resolved after reduction of the doses to 1.2gm per oral stat followed by 0.6gm twice daily. One participant in the severe category B showed prolong QT prolongation on the 3rd day of administration of HCQ and hence the offending drug was discontinued and the trial discontinued for the same patient. Four participants from the non-severe A, three from the non-severe B, and two patients from the severe B had withdrawn from the trial.

**Fig. 1:**
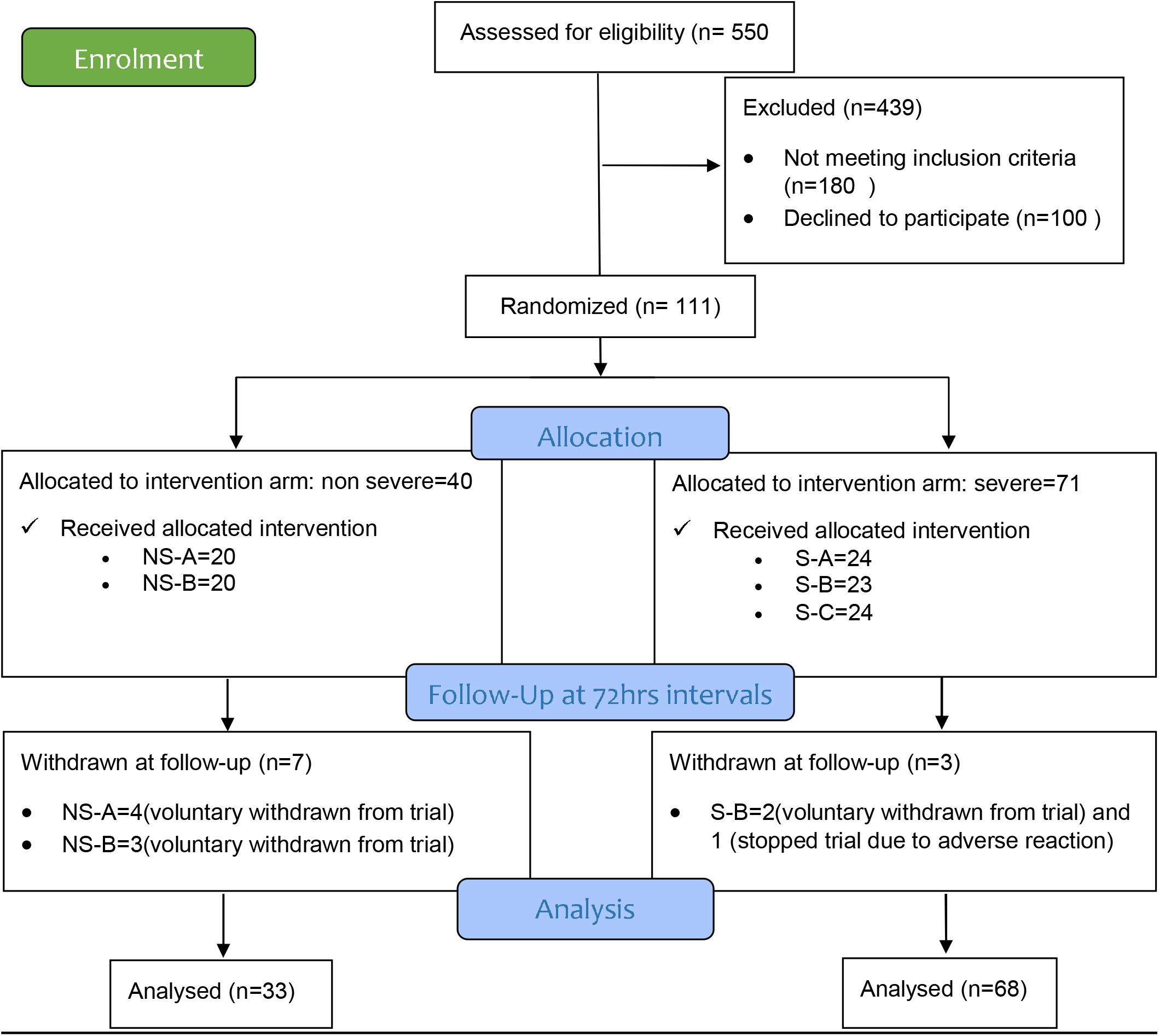
The study flow (NS=Non-severe, S=Severe category of intervention arms)

### Baseline characteristics

In the severe category, cough, shortness of breath, and fever were the primary reasons for participants to present in the health facility (Table 1). Diabetes mellitus, COPD, hypertension, and asthma were the comorbidities with which the participants presented of which hypertension predominated. Similarly, in the non-severe category, participants presented with predominant complaints of fever, cough, and shortness of breath. Also, a trend of male predominance and older age groups were observed in all participants.

**Table 1:**
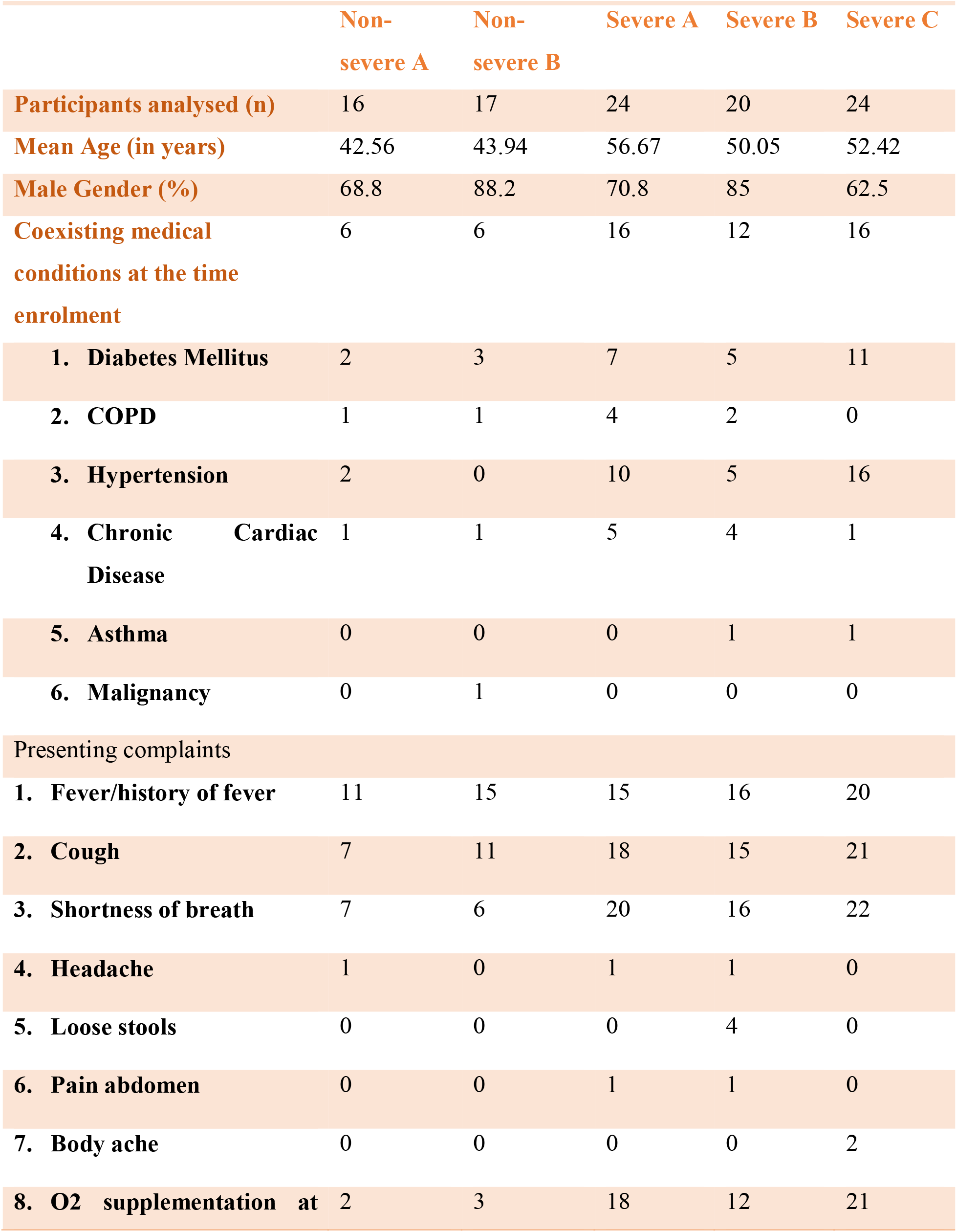
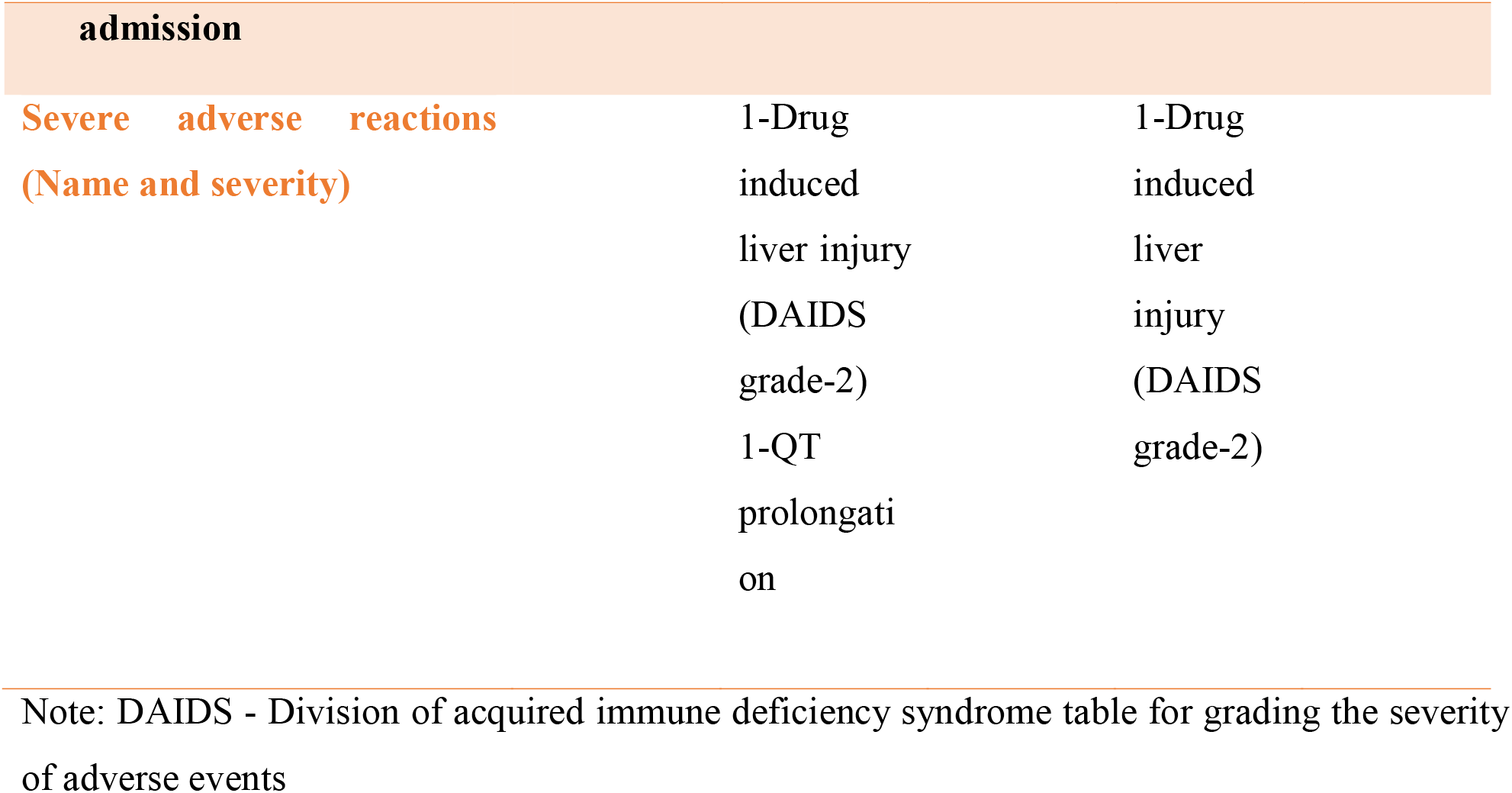
Demographic and clinical characteristics of the patients at baseline.

### Primary outcome

Except for two participants (one from severe B and another from the non-severe B) who developed elevated liver enzymes after high doses of ribavirin (dose reduced to half), and one participant who developed QT prolongation after day 3 of HCQ (the offending agents were stopped), no other participants developed conditions that can be attributed to the trial drugs. The clinical recovery was assessed for presenting symptoms as the depicted trend of resolution of symptoms in the graph (Fig. 2). However, the difference in clinical recovery status was not statistically significant among three groups in the severe category and two groups in the non-severe category.

**Fig. 2:**
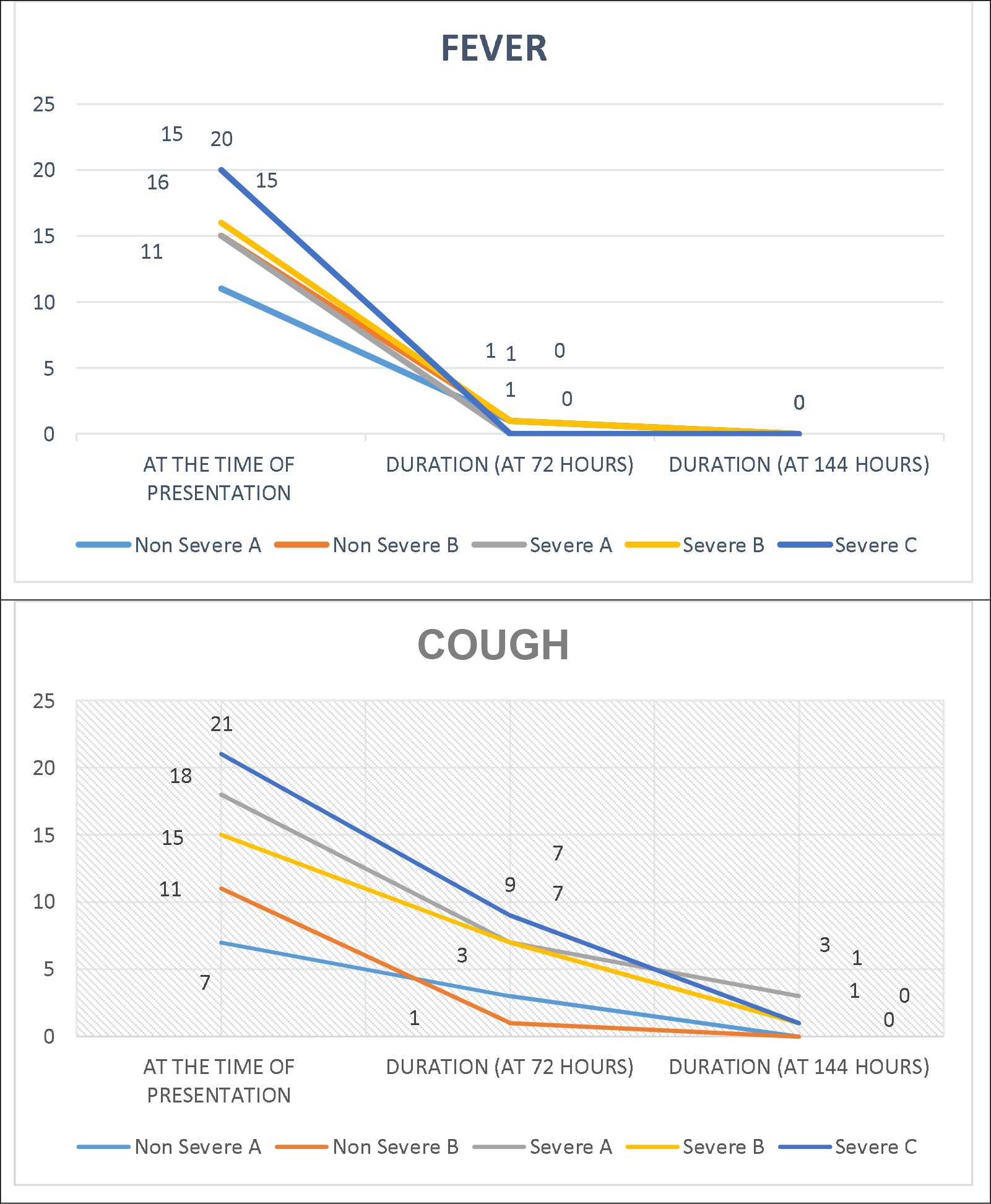
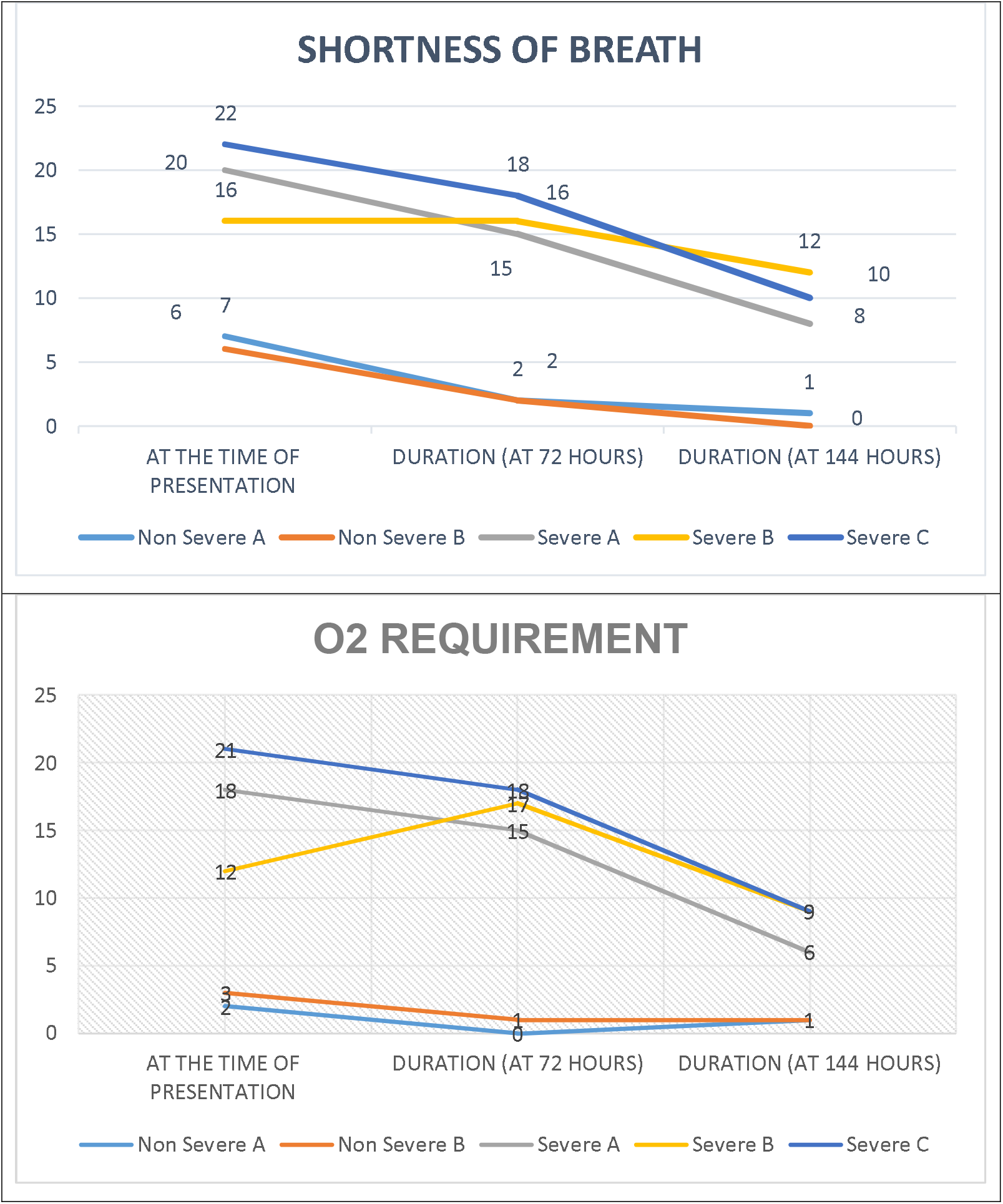
Time trend of clinical and laboratory recovery of major symptoms and tests among different intervention arms Concerning laboratory recovery such as organ function tests (hemogram, liver, and kidney function tests), all groups (both severe and non-severe category) showed similar non-significant increasing or decreasing trends concerning the time of admission and subsequent follow-ups (Fig. 3).

**Fig. 3:**
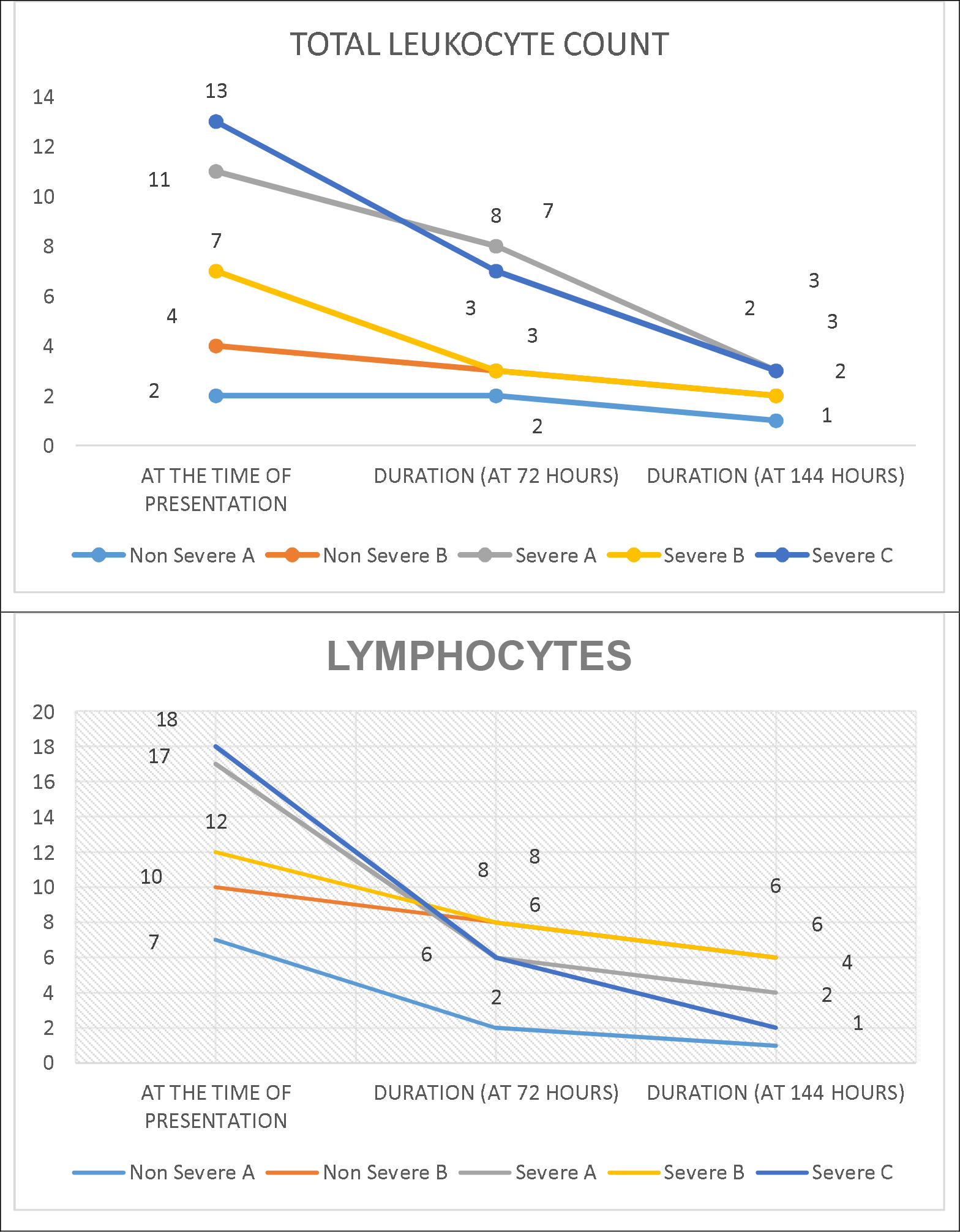
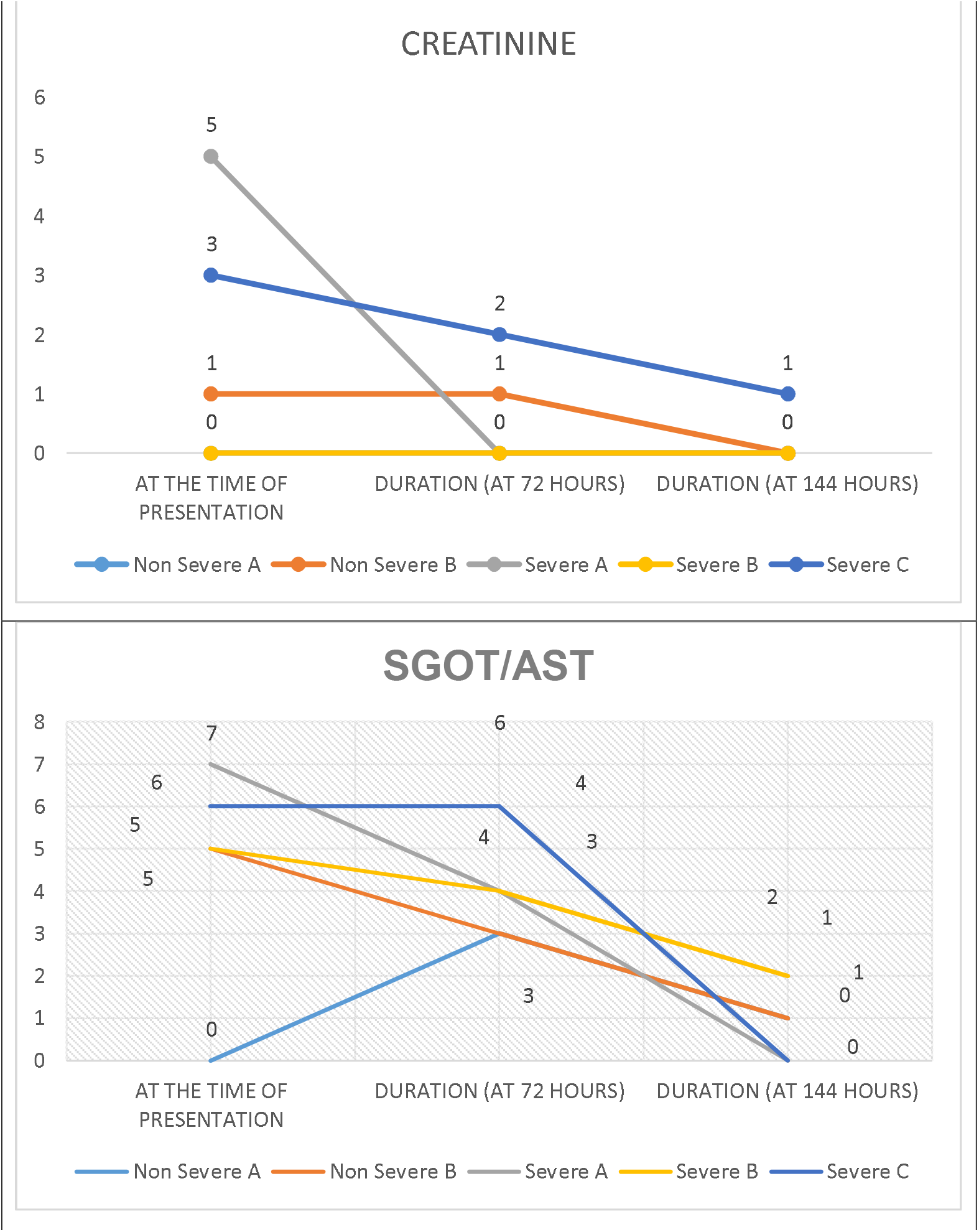
Time trend of laboratory recovery of major tests among different intervention arms The time to SARS-CoV-2 RT-PCR negative in nasopharyngeal swab specimen was also not significant among groups (Table 2).

### Secondary outcome

Analysis of secondary outcomes using the concurrent randomization analysis revealed that all-cause mortality was lowest in category severe B followed by in category severe C and highest in category severe A without any intergroup statistical significance and no mortality was recorded in non-severe categories (Table 3). Similarly, we found no significant increase in the frequency of respiratory progression in the non-severe category, however, in severe categories, the need for positive pressure (both non-invasive and invasive) ventilation at admission gradually declined during consecutive follow-ups in all three groups, least in severe group A, due to inadequate sample size it failed to reflect any statistically significant value. The average duration of hospital stays was two weeks for all severe groups.

**Table 2:** The time to a negative SARS-CoV-2 RT-PCR in nasopharyngeal and throat swab specimense.

| Non- | Non- | Severe | Severe | Severe |
| --- | --- | --- | --- | --- |

|  | severe A | severe B | A | B | C |
| --- | --- | --- | --- | --- | --- |
| No of the participants having Negative RT-PCR at follow-up intervals till discharge | 3 | 3 | 2 | 3 | 2 |
| Mean duration (in days) | 5.66 | 6.33 | 12.5 | 8.5 | 8 |
| P-value | Intergroup >0.05 (non-significant) |  |  |  |  |

**Table 3:**
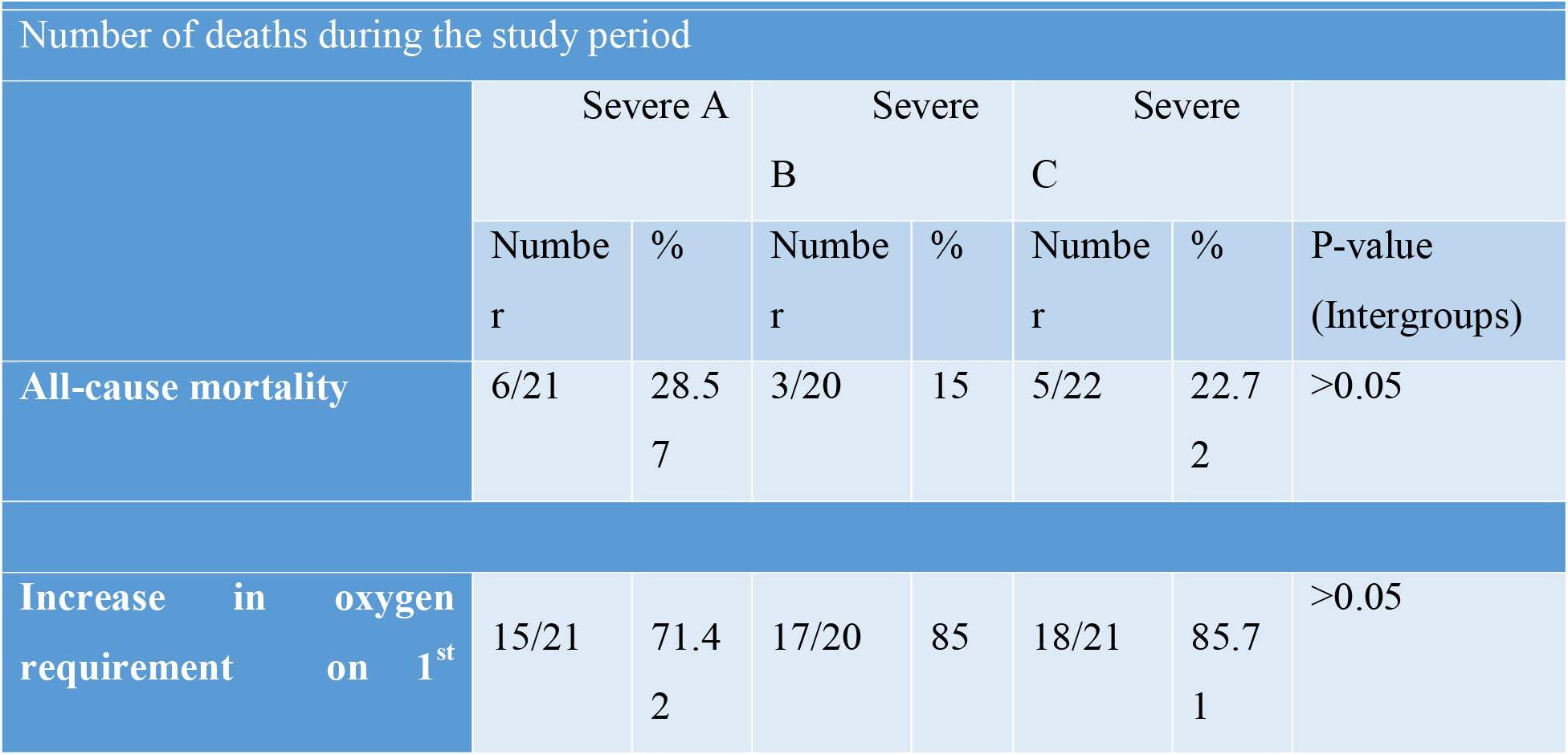
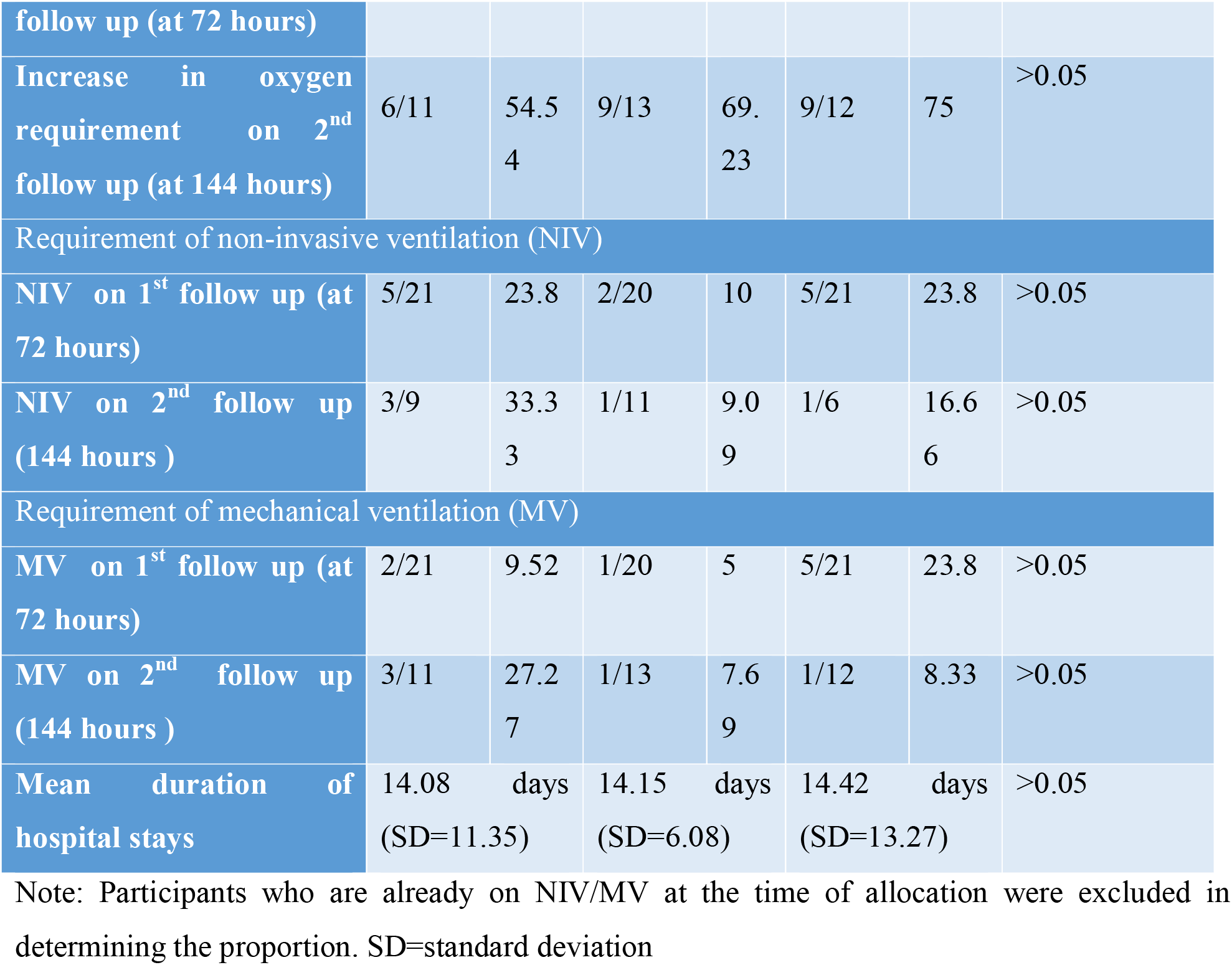
Secondary outcomes among participants in the severe category.

In the case of the non-severe category, no significant finding could be made concerning the trend of increase in oxygen impairments. The duration of hospital stays remained 8.12 days (SD=3.20) in non-severe A and 10.18 days in non-severe B (SD=6.347) respectively. Two participants from non-severe B had the intermittent requirement of NIV, which resolved and recovered in the subsequent follow-ups. One participant in the non-severe B had progression of the disease and was subsequently randomized to the severe groups ultimately leading to the requirement of MV and hence discontinuation of the trial. Two participants from the non-severe A and three participants from non-severe B were supplemented with low flow O2 because of background COPD. By the time of discharge, all participants in the non-severe groups had been weaned off from O2.

## Discussion

This randomized controlled trial attempted to identify antiviral combination therapy for its potential benefits in the treatment of Covid-19 in both non-severe and severe manifestations of the same disease. Although with limited data, no statistically significant findings could be observed.

However, with the available data in hand, certain observations are made that are consistent with the previously published studies involving the antivirals and the disease characteristics. The degree of shortness of breath and other organ dysfunctions were the primary factors that guided in categorizing the severity of COVID-19 infections and this system held as only one patient in the non-severe group progressed to severe symptoms during the trial period with the remaining participants in the category making an uneventful recovery.

In the patient demographics, the predominance of the male sex group in all severities of the study groups is noteworthy. There are also additional findings of concomitant comorbid conditions of which hypertension appears to be the most common among others such as chronic cardiac diseases, COPD, and asthma. This may provide interest in future studies elaborating the intrinsic pathophysiology and disease outcomes concerning COVID-19 among the hypertensives (17)

In the comparative study of the non-severe category between the participants receiving standard treatment and HCQ + ribavirin, nearly all patients showed recovery during the study period and a clear association between the treatment outcomes of the two intervention groups couldn’t be made. Hence, from our study, it can be concluded that in future endeavors towards the management of COVID-19 patients, the decision to treat mild-moderate (non-severe) disease with any form of therapy regardless of their efficacy, it is noteworthy to mention that almost all recover without any difference in the duration of illness or mortality. This may rationalize the use of scarce health care resources during this pandemic (18).

On the other hand, in the severe category, our data also suggest that HCQ + ribavirin have prevented the progression to more severe respiratory disease, as shown by the lower proportion of patients progressing to the need of NIV and MV compared to the standard treatment group without any statistical significance. At the same time, no significant side effects are noted during the study period. Also, all-cause mortality is the lowest in the severe group B. Similar findings are also noted while comparing between the standard treatment group and lopinavir/ritonavir + ribavirin group as no statistically significant difference is noted. The number of samples may have been too short and less to evaluate that subgroup (19).

An amendment was also made in the trial to reduce the doses of ribavirin due to the development of deranged liver enzymes; when this change was implemented six patients had been enrolled in the trial, but only two had received ribavirin in two groups. However, this has given one good outcome that ribavirin shouldn’t be used with 1800mg loading dose and 1200mg BD maintenance dose in the Indian population where it could lead to drug-induced liver injury. This trial establishes lesser ribavirin doses for Indians compared to the western population (20).

This trial did not complete full enrolment owing to the development of new studies suggesting remdesivir to have beneficial effects in O2 requiring patients and also more patients opting for newer agents during the trial period which becomes a major factor in premature discontinuation of the study for those participants (21). When we compare our study results with that of remdesivir, one conclusion that can be drawn is that these non-standard group of treatments can be used for clinical recovery of symptoms and laboratory abnormalities and need of oxygen/ventilator requirements without any mortality benefit similar to remidesivir study. Our study was also non-blinded and lacks heterogeneity of different populations being a single-centered study. With emerging data suggesting COVID-19 to have a more protracted course, different outcomes may have been missed with the limited follow-up period. There are also emerging strains of new COVID-19 with variable degrees of disease manifestations in regards to severity and organ involvement. Hence, concern also arises about the efficacy of these drugs in those newer strains and hence needs further evaluation.

The trial was also not devoid of challenges as it was conducted during a time of restricted travel and restricted entry of nonessential personnel. Monitoring visits often were performed remotely and strict implementation of isolation and other measures of infectious control was a major hindrance in drawing and processing samples in defined periods. There was also a shortage of trial-related supplies in between the study period such as reagents for the inflammatory markers. There was also the publication of results by the WHO from the Solidarity Trial in July 2020, which recommended discontinuation of HCQ and lopinavir/ritonavir arms and this harms enrolment of participants of our study (22).

Given the results of the findings of the study and availability of a variety of therapeutic approaches including novel antivirals, modifiers of the immune response, or other intrinsic pathways, antiviral combinations may have a role in improving clinical recovery in patients of COVID-19 with severe category whereas only standard therapy is required for non-severe category.

## Data Availability

It will be provided with permission from the Corresponding author.

## Trial registration

Clinical Trial Registry of India (CTRI): **CTRI/2020/06/025575, http://www.ctri.nic.in/Clinicaltrials/pmaindet2.php?trialid=43076**

## Ethics approval and consent to participate

This trial was approved by the Institutional Ethics Committee of AIIMS, Rishikesh on 30/05/2020 with reference no. 218/IEC/IM/NF/2020. Written informed consent was obtained from all participants.

## Authors’ contributions

PKP conceived the trial. PKP, AB, BCS, GC, YAB designed the trial. BCS, BM, SS collected the data. All the authors are part of the trial management committee and were involved in the review, amendments, and approval of the final protocol.

## Acknowledgments

Thanks to Prof Manoj Gupta, Prof UB Mishra of the institute for availing logistics required for the trial; to Dr. Arkapal Bandyopadhyay, Dr Ramanuj Samanta, Dr Rohit Walia, Dr Itish Patnaik, Dr Gaurav Chikara, Dr Ravi Gupta, and Dr Bharat Bhusan Bhardwaj, for helping protocol preparation and handling drug complications if any of the study participants; to Dr Deepjyoti Kalita and Dr Puneet Gupta for performing laboratory diagnosis of the study participants.

